# Feasibility, usability and acceptability of a novel digital hybrid-system for reporting of routine health information in Southern Tanzania: A mixed-methods study

**DOI:** 10.1101/2022.02.07.22270225

**Authors:** Regine Unkels, Fatuma Manzi, Ntuli A. Kapologwe, Ulrika Baker, Aziz Ahmad, Rustam Nabiev, Maria Berndtsson, Jitihada Baraka, Claudia Hanson, Atsumi Hirose

## Abstract

**Background:** Health information systems are important for health planning and monitoring of progress. Still, data from health facilities are often of limited quality in Low-and-Middle-Income Countries. Quality deficits are partially rooted in the fact that paper-based documentation is still the norm at facility level, leading to mistakes in summarizing and manual copying. Digitalization of data at facility level would allow automatization of these procedural steps. Here we aimed to evaluate the feasibility, usability and acceptability of a scanning innovation called *Smart Paper Technology* for digital data processing.

**Methods:** We used a mixed-methods design to understand users’ engagement with *Smart Paper Technology* and to identify potential positive and negative effects of this innovation in three health facilities in Southern Tanzania. Eight focus group discussions and 11 in-depth interviews with users were conducted. We quantified time used by health care providers for documentation and patient care using time-motion methods. Thematic analysis was used to analyze qualitative data. Descriptive statistics and multivariable linear models were generated to compare the difference before and after introduction and adjust for confounders.

**Results:** Health care providers and health care managers appreciated the forms’ simple design features and perceived *Smart Paper Technology* as time-saving and easy to use. The time-motion study with 273.3 and 224.0 hours of observations before and after introduction of *Smart Paper Technology*, respectively, confirmed that working time spent on documentation did not increase (27.0% at baseline and 26.4% post-introduction; adjusted p=0.763). Time spent on patient care was not negatively impacted (26.9% at baseline and 37.1% at post-intervention; adjusted p=0.001). Health care providers described positive effects on their accountability for data and service provision relating to the fact that individually signed forms were filled.

**Discussion:** Health care providers perceived *Smart Paper Technology* as feasible, easy to integrate and acceptable in their setting, particularly as it did not add time to documentation.

## Introduction

Quality and timely health information is crucial to strengthen health systems [1, 2] and to monitor achievements of nationally and internationally agreed targets such as the *Sustainable Development Goals* [3]. Although population-based surveys, often done in 5-years intervals only, are the mainstay to generate health data [4], *Health Information Management Systems* (HMIS) data are increasingly valued as they are continuously available and less cost-intensive than surveys [2, 4].

At present, the *District Health Information System* (DHIS2) is used to collect health information in over 70 countries worldwide [5]. Individual patient level data is recorded manually in HMIS paper-based registers, then summarised on monthly report forms in health facilities (figure 1a below). Aggregated summary data from these reports is entered manually in the electronic DHIS2 at district level [6] where various users can view this summary data at different levels of aggregation (figure 1 b below). While the system is well established, incompleteness and inconsistency of data due to calculation and reporting errors have been described [7-10]. Also, digital aggregated data is often not available on time for decision making especially at sub-national level [11]. Another limitation of the present system is the lack of individual-level data making it impossible to construct coverage indicators for sub-groups, such as patients with complications [12].

**Figure 1A:**
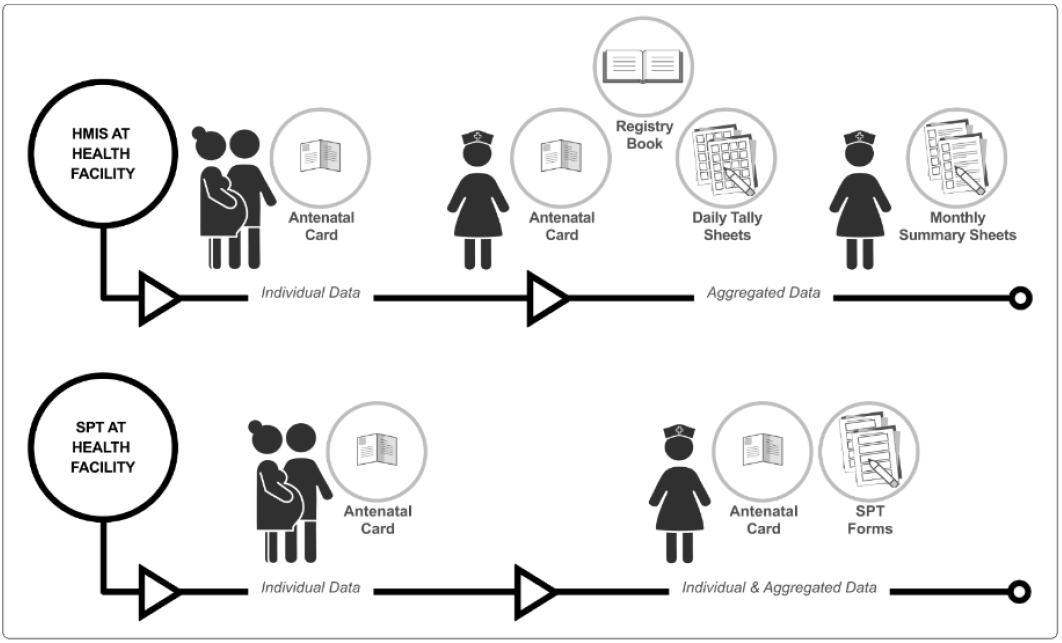
The difference between HMIS and SPT systems at health facility level. <u>HMIS:</u> A nurse documents patient and service information into a woman’s antenatal care card and in a register book. Daily tally sheets and monthly report forms are manually created. The latter are brought to the district headquarter up to 15^th^ of each month. <u>SPT:</u> A nurse documents patient and service information in a woman’s antenatal care card and on one scannable SPT form. Forms are brought to district headquarter in regular intervals.

**Figure 1B:**
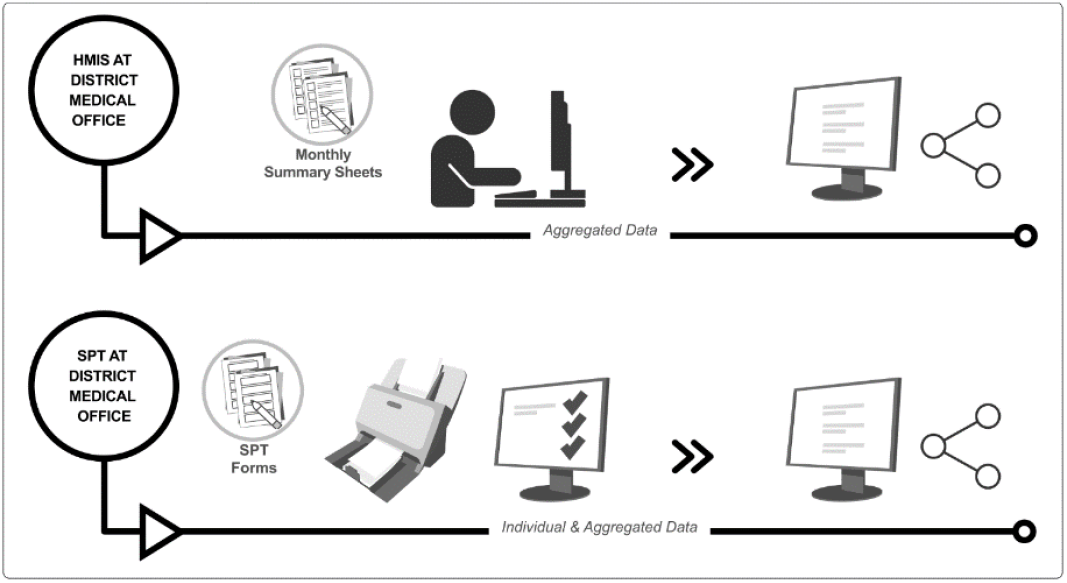
The difference between HMIS and SPT systems at district level. <u>HMIS:</u> Several health care managers manually enter information from monthly report forms of all health facilities into the electronic DHIS2 system. Obvious inconsistencies or errors detected during data entry are followed up by phone or during physical visits where registers are checked. In DHIS2 aggregated data can be viewed from facilities with paper-based data entry (dispensaries) and both individual and aggregated data from fully digitized data entry (many health centers and all hospitals). <u>SPT:</u> One scanning officer scans all forms per facility. Data is automatically read by the software. Inconsistencies or errors are flagged by the system. A data verification officer works on these using the electronic register and phone calls to facilities if needed. Daily tally sheets and monthly summary reports are created automatically. Individual and aggregated data can be viewed.

Digitalization at the first step of a health management information system – where data is generated - has the potential to alleviate these problems [13]. *The Smart Paper Technology* (SPT) system uses an innovative software to scan and digitize information on SPT forms and improve the collection of routine health data (Figures 1a, 1b above). These forms, matching HMIS register content for ante-, intrapartum and postnatal care (Figure 2 Annex), completed at facility level are scanned at district level and generate digital individual patient data as well as aggregated data. In addition to monthly summary reports, the system continuously generates electronic data on a dashboard where both individual and summary data can be viewed. Data inconsistencies are signaled automatically. SPT was first used to support documentation of health data for vaccination services in Uganda and The Gambia [14, 15].

We introduced SPT in one typical district council with 29 primary facilities and one hospital in Southern Tanzania for the continuum of antenatal, intrapartum and postnatal care between May 2019 and June 2020.

Digital innovations need to be relevant to context and user group feedback are important to support implementation and to evaluate potential for scale up [16, 17]. Technology that is not adapted to the context may increase time spent on data generation, negatively affecting service provision and consequently patient safety [18, 19]. End users may fail to integrate the new technology into existing systems with negative effects for sustainability. Taking these recommendations into account, our study aimed to evaluate feasibility, usability and acceptability of SPT to understand why, how and to what extent users engage with SPT and to identify potential positive and negative effects of this newly introduced digital technology.

## Methods

### Overall design and conceptual framework

We adopted a convergent parallel design [20] applying a mix of qualitative and quantitative methods grounded in social constructivism [21]. The PRISM framework (*Performance of Routine Information Management*) developed by Aqil et al. [22] and *Normalization Process Theory* [23] informed our conceptual framework (Figure 3 Annex). *Normalization process Theory* applies four constructs to explain how users integrate new technologies into existing routines and communications: i) coherence (users assign meaning and utility to a new technology) ii) cognitive participation (users engage with new technology by their own agency or by assignment), iii) collective action (users organize and operationalize new technology) and iv) reflexive monitoring (users evaluate new technology informally or via formalized platforms) [23].

Our qualitative study part used focus group discussions (FGDs) with health care providers (HCPs) and in-depth interviews (IDIs) with health care managers exploring feasibility, usability and acceptability after SPT introduction. The quantitative time-motion study compared time spent by HCPs on documentation and patient care before and after SPT introduction.

### Setting

The project was conducted in cooperation with the ministry of health and local government officials at national, regional and district level in one rural pilot district, Tandahimba district council, Southern Tanzania. The district has a population of 200,000 inhabitants, predominantly subsistence farmers [24, 25]. A phased introduction of SPT took place over 13 months, accompanied by mentoring visits by the project team and remote mentoring and troubleshooting via WhatsApp. HCPs were trained on content and completion of forms. Selected managers at district level received training on dashboard use, and towards the end of the 13 months, on data verification. Double data entry into both SPT forms and HMIS registers was requested by the ministry throughout the study to maintain routine reporting to national level.

### Sampling strategy and recruitment

Tandahimba district council was purposefully selected because of a pre-existing research programme [26, 27]. From the 30 facilities within the district council we sampled three public facilities, the only hospital, one health centre and one dispensary to include the three levels of care. Facilities were chosen for accessibility, case load and because they were part of the first scale-up wave.

The qualitative sampling strategy applied purposive sampling for FGDs and IDIs, based on the concept of information power [28], and included participants known to process data for HMIS and SPT for maternal and newborn services.

For the quantitative time-motion study, we included all HCPs from the selected facilities who were present on observation days. Informed by previous studies [29-31], we calculated that 240 hours of observation were needed before and after SPT introduction (after inflating with factor 1.5 to adjust for clustering) to determine a change in working-time spent on documentation (primary outcome) with 80% power at 5% significance level.

### Data collection

Two data collectors fluent in Kiswahili with previous experience in qualitative research (JB, RU) collected qualitative data in July 2019, one month after SPT introduction (“post-intervention”, four FGDs, six IDIs) and in February 2020, eight months after introduction (“follow-up”, four repeat-FGDs, five repeat-IDIs). Informed by the conceptual framework (figure 2 in annex) and literature on evaluation of digitalization in health care and HMIS, we developed topic guides [17, 22, 23, 32] (see topic guides in annex). Tools were pre-tested and further adapted for follow-up data collection to accommodate new themes from first data analysis. FGDs were conducted after working hours and participants travelling from home for this received compensation of approximately 4 US dollar. IDIs and FGDs lasted on average one hour. FGDs were conducted in facilities, in rooms separate from patient care. IDIs were conducted in participants’ offices. Both were audio-taped after written informed consent and field notes were taken. Frequent summaries were provided to participants during interviews to countercheck their information and to enhance trustworthiness.

We collected time-motion data at baseline (February and March 2019) and 2 months after SPT introduction (August 2019). After a 5-days training, two observers per facility with clinical background, shadowed HCPs for their entire shifts, starting from HCPs’ arrival at their consultation desk. Morning and afternoon shifts, as well as weekends, but no night shifts were included. The data collection tool, informed by Pizziferri et al. [33] and initially piloted with health managers, included 141 tasks under 3 categories and 10 sub-categories, to reflect daily tasks of ante-, perinatal and postnatal care (see annex for full list). The tool was translated into Kiswahili, programmed into Open Data Kit (GetODK. Inc, version 1.2.4.1) and administered on tablets. Pilot observations and feedback sessions conducted during observer training reduced inter-observer variability and ensured that tasks were sufficiently defined and discriminated (task calibration) [34].

All data was anonymized and stored on a password-protected computer. We used a protected cloud at Karolinska Institute for data sharing during joint analysis.

### Data analysis

Qualitative recordings were transcribed ad verbatim without review by participants for logistic reasons. We (RU, UB, JB) applied reflexive thematic analysis [35, 36] with initial inductive and later deductive coding in NVivo 12 pro (QSR International) on Kiswahili transcripts. After initial blinded coding of four transcripts, a coding tree was initiated and applied across the data set. Post-intervention and follow-up data sets were analyzed separately to explore changes in feasibility, usability and acceptability throughout the course of the intervention. We then jointly developed sub-categories, categories and themes and applied the conceptual framework for latent analysis. Although we developed themes close to our primary data, they are inspired by the notion of Greenhalgh and Swinglehust that “technology shapes humans but is also shaped by human interaction” [37].

For the time-motion study, we first compared the number and average duration of observed shifts before and after SPT introduction and for potential confounders (level of care, section, cadre & experience) using Fisher exact test. We estimated the *difference in proportion of time spent on individual task categories and sub-categories per average shift* between pre- and post-intervention. We then used multivariable generalised linear models to assess differences in proportion for each task category between pre- and post-SPT implementation adjusting for potential confounders. The category medical doctor was removed from the analysis since there was only one observation for this cadre from the pre-implementation data collection.

### Ethical consideration

We received ethical clearance from the *Institutional Ethical Committee of Ifakara Health Institute* (IHI/RB/No.20 -2018) and *National Institute of Medical Research* (NIMR/HQ/R.8a/Vol.IX/3018) in Tanzania and from the *Ethics Review Board of the Commune of Stockholm* (2019-04022 Gk), Sweden.

We obtained informed written consent from each participant. Observers assessed suitability for observation at each patient encounter according to agreed standards. Patients were informed about the purpose of the observation, the benefits to the population and their right to object.

## Results

We report key results from the qualitative study with eight FGDs and 11 IDIs in total along our conceptual framework. Two men and 16 women participated in FGDs and three women and three men participated in IDIs (see table 1 below). During follow-up a female manager who was previously interviewed was sick. No medical doctor participated in the study but three non-physician clinicians. All other participants were nurse/midwives or assistant nursing cadres.

**Table 1:** Participant characteristics.

| <u>Method</u> | <u>Men</u> | <u>Women</u> | <u>Cadre</u> |
| --- | --- | --- | --- |
| Focus Group Discussions (FGDs) (4 post-intervention, 4 follow-up) | 2 | 16 | Nurse-Midwife 9 |
|  |  |  | Non-Physician Clinician 1 |
|  |  |  | Assistant Nurse 5 |
|  |  |  | Medical Aide 3 |
| In-depth Interviews (IDIs) (6 post-intervention, 5 follow-up) | 3 | 3 | Nurse-Midwife 3 |
|  |  |  | Non-Physician Clinician 3 |

### Qualitative Results: Focus Group Discussions and In-depth Interviews

We identified two themes, seven categories with 20 sub-categories, 30 codes for the post-intervention data set and 65 codes for the follow-up data set (see table 2 in annex).

### Theme 1 Technical Factors Shaping Human Interaction

#### Category 1 Technical factors

During post-implementation data collection, HCPs and managers described substantial benefits from the simple design of forms, automated generation of tally sheets and monthly reports, leading to time-savings. At follow-up, discussion shifted to other less obvious effects of the new system, such as improvements in services delivery and data quality described further below.

> “*Now for HMIS that means one has to enter everything into the system but with this [SPT] it’s straight forward, you just scan. This can reduce the number of people [involved], it will help other people to perform other tasks*.*”* (IDI1_follow-up)

Respondents also reported challenges related to SPT software, e.g. to process corrections on the forms.

> “*Another challenge is that you may have filled the form well, but then you make a mistake e*.*g. 37 instead of 36 and you can’t delete that. You need to start afresh for that very 36 and you have a lot of clients waiting…*.” (FGD3_post-intervention)

In addition, HCP mentioned that paper forms were sent to the district for scanning, so collected data was unavailable for direct use in the facility.

### Theme 2 Human Interaction Shaping the Use of Technology

#### Category 1 Organizational factors

HCPs and managers described how facilities successfully embedded SPT into their daily work. HCPs felt competent using SPT and described creative ways to embed the use of SPT forms into their workflow including i) re-distributing staff, ii) using notebooks or amending registers to track unique identifiers and iii) introducing a folder-based filing system.

> “*We have two rooms for service provision here. In each room we have now put two people. So, if you have made a mistake [in documenting], your colleague [providing services] will see it and teach you*.” (FGD4_post-intervention)

HCPs appreciated joint learning through a WhatsApp group established at the time of first SPT training. Managers valued the increased virtual contact time there for supervision and feedback, circumventing common bottlenecks including time for physical visits and transport issues.

During post-implementation data collection HCPs mainly raised challenges with the new system itself, while conversations during follow-up focused on known health system issues, such as i) lack of human resources, and ii) lack of transport for supervision, distribution and collection of forms. In addition, managers and HCPs mentioned that most HCPs lacked competencies related to data analysis and use.

> “*For this [SPT] we have not yet gone for supervision. Presently we only know that providers do that work because we have seen the report. And also, you can see once someone ran out of forms, they come to ask for new ones. Like this you know, this one uses SPT*.” (IDI3_post-intervention)

Participants were concerned about sustainability and empowerment of managers regarding i) data access, ii) dashboard access and iii) lack of training in data verification and scanning to enable this group to make use of SPT data.

> “*As I have said before we would use it [the data] if we could generate data that is ours. If we could scan, at least we could use them. But for now, to say we have used it is difficult*.*”* (IDI5_follow-up)

#### Category 2 Promotion of Information Culture

Information and data culture was characterized by managers directing HCPs on actions related to data quality and use in a system. The quality of primary data was infrequently checked. Thus, HCPs described their role as rather generating than analyzing data, a perception that was also shared by most managers during interviews.

> *“Charting a graph [from data] is another issue. Because in most cases the data person from the district team is creating to see graphs. For us in the facility, to say we are able to prepare our data or a report ourselves, like creating a graph or doing the analysis, we can’t do that*.*”* (FGD3_post-intervention)

Although managers, especially those working directly with HMIS data, valued data quality highly for this system, they paid less attention to quality of primary SPT data entry, e.g. by providing physical supervision to directly monitor data entry.

> “*Oh yes, these days it’s about data. Every work we do, we must use data. So, data is very important for our work and this is why we emphasize to them [HCP] that what they report needs to be of quality*.” (IDI3_follow-up)

#### Category 3 Behavioral Factors

HCPs explained how use of SPT forms changed their perceptions of service documentation from being a burden and easy to falsify (“to cook up”) for HMIS, to acknowledging the importance of entering correct data for SPT.

> “*This shows straight who filled the form because you must sign*…*That SPT is easy to use because of its transparency. It shows who completed it. But those registers… if something happens you start to search*… *And in some, pages are torn or gone. It is easy to* ***cook up*** *that information*. “(FGD1_follow-up)

At follow-up, HCPs talked about positive effects of SPT on their efforts to provide good quality documentation. While they described how easy it was to omit or enter made-up data in HMIS registers, because these were hardly followed-up on, HCPs reported a different view on data entry for the SPT system. The fact that a signature and name are required on the form prompted them to leave no box blank and to report truthfully.

#### Category 4 System Processes

HCPs developed measures to ensure completeness: HCPs i) reminded each other to complete forms, ii) instructed those who made mistakes and iii) taught newly posted HCPs how to complete and check SPT forms. Operational challenges included consistent availability of SPT forms. Stock-out at district level occurred several times during the evaluation and HCPs reported about the difficulties to catch up with data entry once new forms arrived. Facilities established various ways to achieve constant supply of forms, e.g. by using other planned trips to the district capital to collect new forms. However, a common way to avoid stock-out was borrowing forms from near-by facilities.

> “*That [stock-out of SPT forms] has happened. I know that in the smaller facilities they were finished because some [HCPs] came here to borrow, but here we didn’t have problems, we are receiving them [forms] timely*.*”* (FGD4_follow-up)

HCPs described new quality assurance strategies for SPT, such as i) individual checks against HMIS register, ii) peer-check at shift end and iii) checks by immediate superior, which they had not applied to HMIS data previously.

> “*Besides that [check by facility in-charge] we do our own check-up: If today I have seen six patients, my SPT forms should match that number*… *If they do not match, you must start checking which patient you omitted and how*.” (FGD3_follow-up)

Managers continued to use the same system of feedback, learning and quality assurance applied to HMIS data at their level and did not carry out additional supervision visits for SPT. Both managers and HCPs mentioned however, that the new WhatsApp group increased exchange and feedback regarding data, albeit for SPT only.

> *“Feedback… has increased [with SPT] because with HMIS this is until someone [from the district team] comes and gives you feedback…, and this is not easy. Because you just take your report there, other information they prepare on their own [at district level] to look at all facilities that are doing well and how they are doing*.*”* (FGD1_follow-up)

#### Category 5 Output

HCPs initially raised issues with the design of the form and stockouts as main reasons for impaired data quality. Reflections during the later implementation period shifted to lack of human resources and workload as main obstacles to completing forms correctly

Despite these limitations, HCPs perceived that SPT led to better quality data. Managers were less convinced but still saw opportunities for data quality improvement once double data entry in SPT forms and HMIS registers was no longer required by the ministry of health.

HCPs stated they were confident about the system they had developed to ensure good SPT data collection and data completeness, despite persisting discrepancies between reports.

> “*… Once I have completed my form well, I put all information in the HMIS register and thus at the end of the month, I don’t get into trouble because it [SPT] is as if I had already tallied*.” (FGD2_follow-up)

However, when comparing the reports generated from both systems, a discrepancy in data was observed. Managers dismissed HCPs’ claims regarding lack of human resources but rather stated HCPs’ i) lack of commitment, ii) forgetfulness, iii) double data entry as main reasons for this discrepancy but did not offer solutions on how to improve this.

> *“It is possible… that sometimes, someone forgets that she registered here and forgot there*.” (IDI2_post-intervention)

Nevertheless, all participants reported to be convinced that SPT use alone, without duplicate data entry, would lead to greater data completeness and expressed a hope that SPT would be approved by the ministry of health.

#### Category 6 Outcome

HCPs described that their efforts to integrate SPT in their daily work, led to re-organization of service provision and teamwork. Several HCPs described how SPT use promoted accountability for service provision and indirectly also improved their working environment. HCPs argued that before SPT, they had little incentive to do that.

> “*So, we have organized ourselves well to ensure that everything needed for SPT or maternal and child health is available. We had these BP machines, those manual ones, but now we have automatic, Urine [sticks] are available, HB, here is the syphilis [test]. So, we make sure everything is there to provide relevant services at the right time so things can move forward*. “(FGD2_follow-up)

Managers noted the opportunity for increased accountability, because they could potentially see if services were provided or not and by whom.

> “*The change we noted is that with this SPT you will identify service providers quickly, because if they don’t provide services, they won’t bring the forms*.” (IDI1_follow-up)

#### Quantitative results: Time-Motion-Study

We observed a total of 273.3 hours before and 224.0 hours after SPT introduction in the three pilot facilities. A total of 3,354 tasks was observed pre-and 3,803 post-intervention. An average observed shift ranged from 0.3 hours to a maximum of 19.7 hours (median (IQR) 5.4 (4.4 – 6.1) hours), with no differences between baseline (median (IQR) 5.5 (4.0 – 6.0) hours) and follow-up assessment (median (IQR) 5.3 (4.4 – 6.1) min) (table 3 in annex).

We saw no difference in the median duration of observed shifts across level of care and health care cadre during pre- and post-intervention assessments. We documented, however, more antenatal care services during baseline in comparison to post-intervention.

We observed that around 10% of time of an average shift was spent on *HMIS documentation*: 11.6% (95% CI 7.7, 15.5) before and 9.8%, (95% CI 7.4, 12.1) after SPT implementation (p 0.627) (table 4). *SPT documentation* only added 3% of time spent on *overall documentation* during an average shift (CI 1.9, 4.1). The proportion of time spent on *overall documentation* per average shift did not differ (27.0% (CI 22.6, 31.4) and 26.4% (CI 22.7, 30.7); adjusted p=0.763) (table 4 below).

**Table 4:**
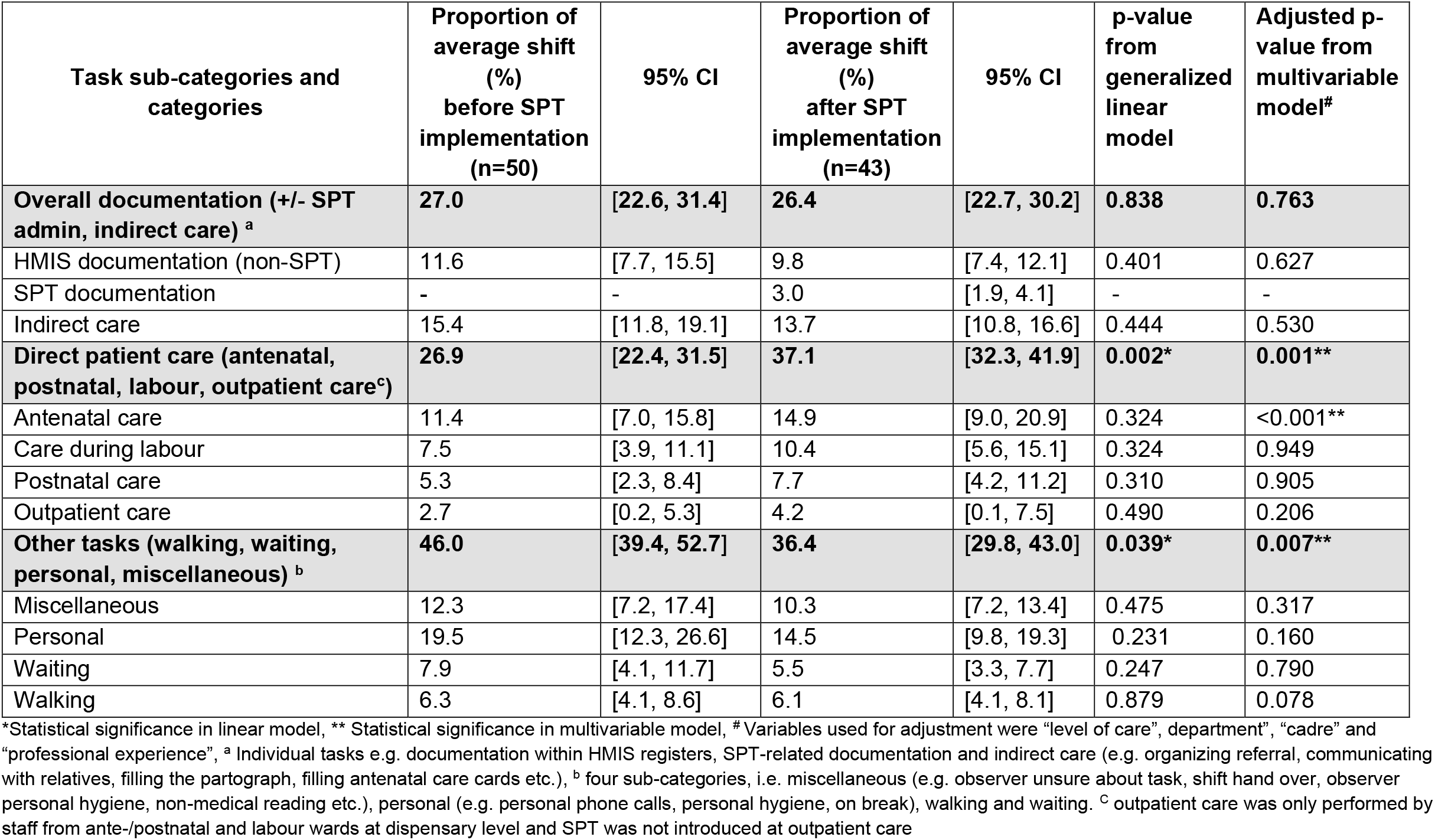
Proportion of time spent per average shift by task sub-categories and categories.

| Task sub-categories and categories | Proportion of average shift (%) before SPT implementation (n=50) | 95% CI | Proportion of average shift (%) after SPT implementation (n=43) | 95% CI | p-value from generalized linear model | Adjusted p-value from multivariable model<sup>#</sup> |
| --- | --- | --- | --- | --- | --- | --- |
| **Overall documentation (+/- SPT admin, indirect care) <sup>a</sup>** | **27.0** | **[22.6, 31.4]** | **26.4** | **[22.7, 30.2]** | **0.838** | **0.763** |
| HMIS documentation (non-SPT) | 11.6 | [7.7, 15.5] | 9.8 | [7.4, 12.1] | 0.401 | 0.627 |
| SPT documentation | - | - | 3.0 | [1.9, 4.1] | - | - |
| Indirect care | 15.4 | [11.8, 19.1] | 13.7 | [10.8, 16.6] | 0.444 | 0.530 |
| **Direct patient care (antenatal, postnatal, labour, outpatient care<sup>c</sup>)** | **26.9** | **[22.4, 31.5]** | **37.1** | **[32.3, 41.9]** | **0.002\*** | **0.001\*\*** |
| Antenatal care | 11.4 | [7.0, 15.8] | 14.9 | [9.0, 20.9] | 0.324 | <0.001** |
| Care during labour | 7.5 | [3.9, 11.1] | 10.4 | [5.6, 15.1] | 0.324 | 0.949 |
| Postnatal care | 5.3 | [2.3, 8.4] | 7.7 | [4.2, 11.2] | 0.310 | 0.905 |
| Outpatient care | 2.7 | [0.2, 5.3] | 4.2 | [0.1, 7.5] | 0.490 | 0.206 |
| **Other tasks (walking, waiting, personal, miscellaneous) <sup>b</sup>** | **46.0** | **[39.4, 52.7]** | **36.4** | **[29.8, 43.0]** | **0.039\*** | **0.007\*\*** |
| Miscellaneous | 12.3 | [7.2, 17.4] | 10.3 | [7.2, 13.4] | 0.475 | 0.317 |
| Personal | 19.5 | [12.3, 26.6] | 14.5 | [9.8, 19.3] | 0.231 | 0.160 |
| Waiting | 7.9 | [4.1, 11.7] | 5.5 | [3.3, 7.7] | 0.247 | 0.790 |
| Walking | 6.3 | [4.1, 8.6] | 6.1 | [4.1, 8.1] | 0.879 | 0.078 |

\*Statistical significance in linear model, \*\* Statistical significance in multivariable model, <sup>#</sup> Variables used for adjustment were “level of care”, department”, “cadre” and “professional experience”, <sup>a</sup> Individual tasks e.g. documentation within HMIS registers, SPT-related documentation and indirect care (e.g. organizing referral, communicating with relatives, filling the partograph, filling antenatal care cards etc.), <sup>b</sup> four sub-categories, i.e. miscellaneous (e.g. observer unsure about task, shift hand over, observer personal hygiene, non-medical reading etc.), personal (e.g. personal phone calls, personal hygiene, on break), walking and waiting. <sup>c</sup> outpatient care was only performed by staff from ante-/postnatal and labour wards at dispensary level and SPT was not introduced at outpatient care

We observed that time spent in the aggregated category *direct patient care* increased from 26.9% (22.4, 31.5) per average shift at baseline to 37.1 % (CI 32.3, 41.9) at post-intervention (adjusted p 0.001) (table 3). We saw no significant change in time per average shift spent on individual task sub-categories related to *patient care* with the exception of *antenatal care*. Here we noted an increase in proportion of time spent from 11.4% (CI 7.0, 15.8) to 14.9% per average shift (CI 9.0, 20.9) after adjustment (p <0.001) (table 4 below).

We observed a significant change in the aggregated category *other tasks*, which decreased from 46.0% (CI 39.4, 52.7) to 36.4% (CI 29.8, 43.0) (adjusted p 0.007) per average shift after SPT introduction. However, changes within the respective sub-categories did not reach statistical significance (table 4 below).

## Discussion

Our findings indicate that HCPs and managers perceived SPT as time saving, functional and well-aligned to pre-existing work- and documentation-processes supporting its acceptability and usefulness within the Tanzanian context. Results from the time-motion study indicated no significant difference in time spent on documentation of an average shift before and after its introduction (27.0% to 26.4%; adjusted p 0.763) and no negative impact on time spent on patient care (26.9% before, 37.1% after SPT introduction; adjusted p=0.001) per average shift. These findings confirm our qualitative results where respondents described the system as time-saving due to the simplicity of the form design and due to service re-organization. Our quantitative findings do not support qualitative findings from HCP interviews where participants talk about their workload as a reason for not completing SPT forms for all clients since time spent on other tasks such as private tasks took up 46.0% per average shift before and 36.4% per average shift after SPT introduction.

HCPs described positive effects on feeling accountable for data quality and service provision driven by the new tool.

Our findings that SPT was well accepted by users for format and layout leading to time-savings was also raised by two studies evaluating SPT for child vaccination services in Uganda and The Gambia [31, 38]. However, other studies reporting on various other digital technologies such as electronic patient records also indicate high satisfaction with electronic tools, a finding that may suggest that HCPs generally accept digital technology well [29, 39].

We used Normalization Process Theory to analyze feasibility and acceptability of embedding SPT in the workflow: HCPs identified benefits for their work early after SPT introduction (*coherence*). HCPs used re-organization of documentation and service delivery processes to embed SPT into their work but managers applied already established support strategies, e.g. physical supervision and feedback via phone calls (*cognitive participation*). Although the WhatsApp group could have been used as a platform for *reflexive monitoring* and *collective action*, HCPs’ perceived inability to analyze and use data prevented this and participants carried on with the established system of HCPs only receiving information from analysis performed by managers.

A study reporting on the integration of an online-community into clinical routine in the Netherlands, used NPT to highlight how HCPs developed strategies to integrate this tool into current work, similar to our participants, e.g. taking a dedicated time to answer questions from the forum [40].

We report no difference in time spent on overall documentation after SPT introduction despite double data recording using the new SPT while continuing with the present HMIS. The ministry of health asked to maintain data entry into HMIS during the evaluation to keep up national reporting of essential indicators and as a reference system for the evaluation of SPT data quality and completeness. In view that time spent on SPT documentation was only 3.0% per average shift, whereas time spent on HMIS documentation was 9.8% per average shift after SPT introduction, we believe that SPT once introduced as the only documentation system could be a time saver.

A study evaluating SPT for vaccination services in Uganda found a neglectable 24 seconds increase in documentation time per immunization service [31]. Another SPT evaluation of vaccination services in The Gambia found a reduction by 16 minutes per child [38]. In contrast, evaluations using other options than SPT pointed to an increased time spent on documentation. Were et al. using time-motion studies to evaluate introduction of electronic patient records in Uganda, reported increased waiting time for patients and reduced time spent with clients. Authors argued that this finding may be due to HCPs being unfamiliar with the digital technology and thus taking more time for documentation after clients had left [29]. Similarly, Rotich et al. described reduced contact time with patients and increased time for patient registration after the introduction of electronic patient records in a rural health center in Kenya [30].

We report that the simple fact that SPT forms contain HCPs’ name and signature, increased perceived accountability. It is surprising that a design component of the form impacted accountability in this way. Although low numbers of HCPs in the district could have also associated individuals to HMIS data previously, our participants did not perceive this as a trigger for responsibility towards HMIS data correctness in the same way. Other publications [41, 42] including the PRISM framework place accountability under the subject of organizational culture [22]. Our findings propose that a design feature – the name and signature – can affect accountability. This finding warrants further exploration.

We used the PRISM framework as our basic conceptual model which was developed as a quantitative tool to assess the performance of *existing* routine health information systems. We argue that a qualitative context exploration including the prevailing information culture and emerging adaptations through user interaction with the technology would be crucial to document implementation of a *new* system. Consequently, we added *Normalization Process Theory* to understand these processes i.e. how HCPs made sense of SPT and collectively integrated it into their existing work, which is important given the context this took place in (Figure 4 below). This model also supported us to generate a theory on how the technology may have shaped human (inter)action and how this interaction may have shaped the use of the very technology to impact processes, outputs and outcome.

**Figure 4:**
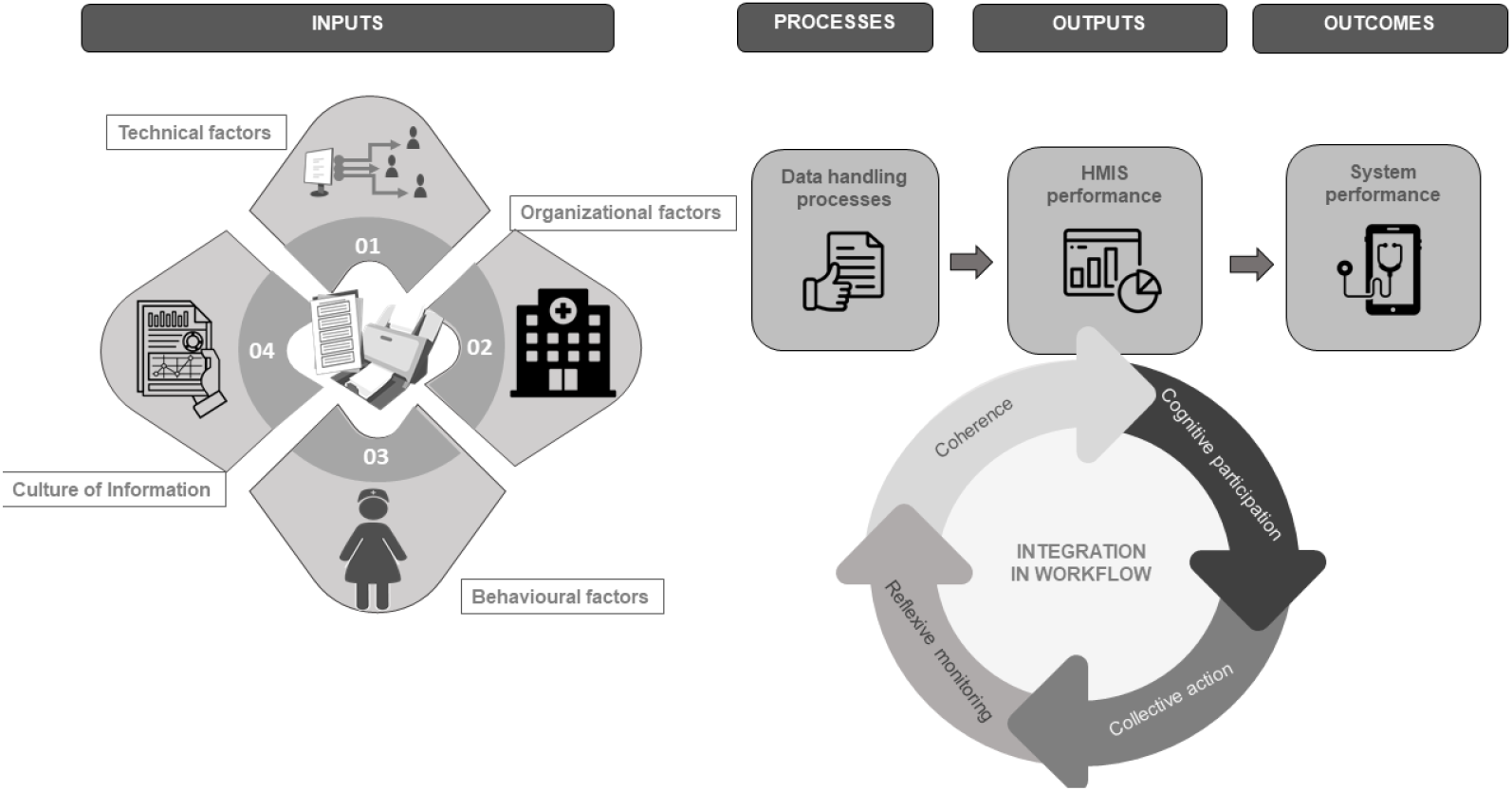
Factors influencing the use of digital technology to impact documentation processes, outputs and outcome.

Our mixed-methods design has several strengths, allowing for triangulation, which is distinctly different from most other studies assessing user acceptance, applying quantitative surveys alone or in combination with other quantitative methods such time-motion studies [29]. To our knowledge there is only one peer-reviewed study that combined in-depth interviews with time motion methodology to evaluate aspects of usability, feasibility and acceptability of a new digital technology in similar settings, though health managers perception was not explored [31]. In addition, our design and conceptual framework allowed the evaluation of the entire system of health data processing and its determining factors.

However, our study also faces limitations. First, we only conducted the time-motion study in three out of 30 health facilities. The observed change in percentage of time spent on documentation was much smaller than anticipated, reducing the power of the study. The study did not include any investigation of the time spent on further processing of data at district level, thus we did not compare health managers’ time spent on documentation. No data was collected on patient load during the two periods of observation as a potential confounder.

Our Observers changed between data collections due to other commitments of the first group. This may have influenced the number of individual tasks recorded due to different experience of both groups. However, we kept inter-observer variability low by including practice observations with feedback as described in our methods section and by Zheng et al. [34]. Observation may have introduced a Hawthorne effect, possibly prompting HCPs to increase their efforts related to documentation but also with regards to patient care.

Our results indicate that it is feasible to integrate SPT in resource-constraint settings such as rural Tanzania with a high degree of acceptability among users and without negative impact on overall documentation and patient care. Our evaluation indicated that workflow was improved through service re-organization, which in turn let to increased service provision and also, interestingly, to improved services through accountability mechanisms triggered by the design of the SPT form itself. It seems important to document similar collateral effects during the introduction of digital tools.

## Supporting information

ICMJ disclosure form

## Data Availability

All quantitative data are contained in the manuscript or supplementary material.
No raw data for the qualitative study part will be made available because it relates to a small, specific participants group in three health facilities in one district in Tanzania. Making the full data set publicly available could potentially breach the privacy that was promised to participants when they agreed to take part and the ethics approval granted from the National Institute for Medical Research in Tanzania and the Swedish Ethical Review Authority. Therefore, the authors will not make the full transcripts available to a wider audience. However, a summary table of codes, categories and themes is provided in the supplementary material.

## Abbreviations

DHIS2: District Health Information System 2
FGD: Focus Group Discussions
IDI: In-depth Interviews
HCP: Health Care Provider
HMIS: Health Information Management System
PRISM: Performance of Routine Health Information Systems Management
SPT: Smart Paper Technology

## Acknowledgement

We would like to thank all the health care providers and managers who took the time to share their thoughts and experiences with us and agreed to be followed during the time-motion study. We are also tremendously grateful to the clients who agreed to their consultation being observed and to regional and district authorities for allowing us to conduct this research.

