## Supplementary material for "Feasibility, usability and acceptability of a novel digital hybrid-system for reporting of routine health information in Southern Tanzania: A mixed-methods study": Figure 2

### REGISTRATION FORM

Health facility DEMO    Date day - month

01 1 02

Identification number **000-0092**

Name ______ Partner name ______

Age ______ Height (cm) ______ Local government chairperson ______

Village ______ Street or/Sub-village ______ Phone number ______

***Pregnancy history***

Previous CS ◯ Age of youngest child ______

***Number of:***

Miscarriages/Abortions ______ Newborn deaths (first week) ______

Live births ______ Stillbirths ______ Children alive ______

***Current pregnancy***

Duration of pregnancy (weeks) ______

day - month

Date of last menstrual period ______ - ______

Estimated date of delivery ______ - ______

Identification number **000-0101**

Name ______ Partner name ______

Age ______ Height (cm) ______ Local government chairperson ______

Village ______ Street or/Sub-village ______ Phone number ______

***Pregnancy history***

Previous CS ◯ Age of youngest child ______

***Number of:***

Miscarriages/Abortions ______ Newborn deaths (first week) ______

Live births ______ Stillbirths ______ Children alive ______

***Current pregnancy***

Duration of pregnancy (weeks) ______

day - month

Date of last menstrual period ______ - ______

Estimated date of delivery ______ - ______

01 1 02



DEMO

### REGISTRATION FORM

01 2 02

Identification number **000-0117**

Name | Partner name

Age | Height (cm) | Local government chairperson

Village | Street or/Sub-village | Phone number

***Pregnancy history***

Previous CS ○ | Age of youngest child

***Number of:***

Miscarriages/Abortions | Newborn deaths (first week)

Live births | Stillbirths | Children alive

***Current pregnancy***

Duration of pregnancy (weeks)

day – month

Date of last menstrual period –

Estimated date of delivery –

Identification number **000-0125**

Name | Partner name

Age | Height (cm) | Local government chairperson

Village | Street or/Sub-village | Phone number

***Pregnancy history***

Previous CS ○ | Age of youngest child

***Number of:***

Miscarriages/Abortions | Newborn deaths (first week)

Live births | Stillbirths | Children alive

***Current pregnancy***

Duration of pregnancy (weeks)

day – month

Date of last menstrual period –

Estimated date of delivery –

01 2 02

DEMO
