## Supporting tables, figures and material for "Feasibility, usability and acceptability of a novel digital hybrid-system for reporting of routine health information in Southern Tanzania: A mixed-methods study"

#### Annex

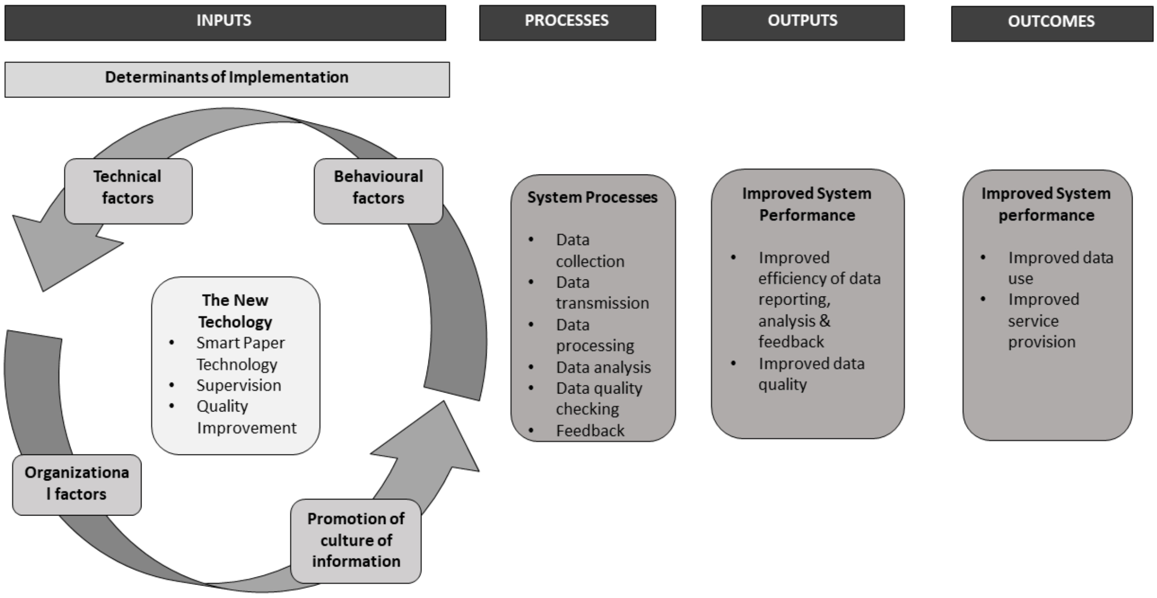

**Figure 3: Conceptual framework of the study**

**Table 2: Overview of identified themes and categories**

| **Themes** | **Categories (informed by PRISM)** | **Sub-Categories** |
|---|---|---|
| Technical factors shaping human interaction | Technical factors | Challenges with SPT system use |
| | | Benefits of SPT system design |
| | | Challenges with existing system |
| Human interaction shaping the use of technology | Organizational factors | Contextual challenges affecting SPT use |
| | | Challenges associated to lack of training |
| | | Support systems already in place |
| | | Responsiveness to innovation |
| | | Support of facility leadership to SPT |
| | | District managers' support to Institutionalizing SPT |
| | Promotion of information culture | HCPs not empowered to use data |
| | | Managers' views on data |

| | | |
|---|---|---|
| | Behavioral factors | Challenges with abilities to analyze data |
| | | HCPs support implementation of SPT |
| | | HCPs' views on data |
| | | Expectations |
| | System processes | HCPs supporting each other to implement SPT |
| | | Feedback to support data quality and use |
| | Outputs | Strengthened accountability |
| | | Potential use of SPT data |
| | Outcome | Improved HCP performance |

**Table 3: Distribution of shifts observed before/after SPT introduction**

| Potential confounding variables | | Number of observed shifts | | p-value ⁺ | Median observed duration in min by shift (Interquartile range) | |
|---|---|---|---|---|---|---|
| | | **pre- SPT intervention (n= 50)** | **post- SPT intervention (n= 43)** | | **pre- SPT intervention** | **post- SPT intervention** |
| **Level of care** | Dispensary | 9 | 7 | 0.651 | 144.9 (126.3 – 154.3) | 145.8 (117.0 – 176.7) |
| | Health centre | 17 | 11 | | 332.9 (263.9 – 392.3) | 333.7 (289.2 – 390.2) |
| | District hospital | 24 | 25 | | 336.0 (301.6 – 364.8) | 337.5 (305.1 – 363.5) |
| **Department** | Antenatal care clinic | 29 | 12 | 0.005* | 309.2 (154.3 – 364.2) | 297.1 (212.2 – 367.2) |
| | Labour ward | 15 | 16 | | 332.9 (291.8 – 350.6) | 339.7 (321.5 – 379.5) |
| | Postnatal care ward | 6 | 11 | | 325.4 (271.2 – 365.4) | 310.7 (286.9 – 351.1) |
| **Cadre** | Nursing diploma | 25 | 26 | 0.498 | 333.5 (293.9 – 395.2) | 339.7 (310.7 – 381.2) |
| | Nursing certificate | 20 | 15 | | 230.9 (143.4 – 327.5) | 255.2 (145.9 – 307.7) |
| | Other | 5 | 2 | | 351.2 (346.3 – 356.8) | 317.6 (264.2 – 370.9) |
| **Professional experience** | 1-2 years | 22 | 13 | 0.233 | 269.6 (145.1 – 346.3) | 197.8 (145.9 – 289.2) |
| | 3-4 years | 26 | 25 | | 332.9 (277.0 – 364.2) | 339.1 (311.4 – 363.5) |
| | > 5 years | 2 | 5 | | 810.8 (438.4 – 1,183.2) | 310.7 (305.1 – 370.9) |

| | | | | | |
|---|---|---|---|---|---|
| **Total** | 50 | 43 | | 327.5 (239.8 – 362.1) | 317.2 (264.2 – 363.5) |

* Statistical significance, + Fisher exact test

**Annex A: Topic guide focus group discussion post-implementation (English version)**

### TOPIC GUIDES FOR REPEAT FOCUS GROUP DISCUSSIONS (FGDS) WITH HEALTH WORKERS

#### Respondents

Health workers involved in maternal and newborn care provision in intervention facilities

#### Objective

To establish the acceptability, adoption, feasibility and appropriateness of the adapted Shifo Smart Paper Technology ™ system for maternal and newborn care and to evaluate its usefulness for supporting ongoing quality improvement activities in health facilities.

#### INTRODUCTION

*Purpose of the interview*

We are in the process of testing a new and simplified health information system in health facilities in this district. We would therefore like to understand how health workers use the current health information system (HMIS) in this health facility, what other /parallel/ reporting systems may be in place and later to return to learn about your experiences of the new system once it has been implemented.

*Introduce the interviewer and translator*

*Consent procedure*

*Check-list demographics (same for all three repeat FGDs)*

#### **SECTION 2:** SECOND FGD
#### EARLY ASSESSMENT OF SHIFO TECHNOLOGY IMPLEMENTATION

##### INTRODUCTION

*Last time we met in this group, we discussed about the way you are documenting and using health information for mothers and newborns in your health facilities.* ***(Possibility: To provide a summary of what was discussed during the previous FGD; this can serve both as a reminder and starting point of this new FGD as well as a way of "validating" and feeding back the results from the first FGD).*** *Since then, the project has introduced the new system which has changed the processes around documenting, reporting and feed-back of health information. We are now interested to learn about your experiences of this new system and how you perceive it compared to the regular HMIS system, what is good and what is challenging and whether it has changed the way you use health information.*

1A. **What did you think when you first heard about the smart paper technology during training?**

(Prompts: *How difficult was it to understand? What were your feelings about how to apply the new forms? What did you think when you first used SPT? How difficult was it to understand, or to use? How long did you need to get used to filling the forms? How did others in your department perceive the complexity of the new forms? When the SPT technology was introduced to you for the first time, what did you think, the new practice would do for you or change in your work? Have these changes materialized?)*

1B. **How did your superiors and colleagues support you in using the SPT forms?**

(Prompts*: How did they make sure that you have everything you need in terms of knowledge and equipment to use the new system? How do you normally make sure that a new tool or a new tsk is implemented well in your service? Is there a difference to how you did it with the SPT forms? If yes how? How did you organize yourselves in the department to make sure that the SPT system is operating well? How did you make sure that you are all on the same level of knowledge and skills to use the form well?* )

1 C. **Can you describe any changes you have experiences in the *processes* of documenting health information for mothers and newborns in your health facilities following the introduction of the SPT system**

*(Prompts: Where and when do you document, ave any challenges you experienced with the regular HMIS system been solved by the new system?)*

1 D. **Can you describe any changes you have experiences in the *monthly reporting* of health information for mothers and newborns in your health facilities following the introduction of the SPT system**

*(Prompts: How is it different, who is responsible, time taken, when is it done, have any challenges you experienced with the regular SPT system been solved by the new system?)*

1 E. **How has the new system affected your daily work (positive, negative, not at all) with documenting care for mothers and newborns in the health facility?**

*(Prompts: How do you perceive the effects, the new system has on your general work in your department? How do you perceive the effect of the new system on your work as a team? Which changes do you like, which changes do you dislike? How do you perceive the amount of time spent on patient documentation or retrieval of information with the new system as compared to the HMIS book? Which factors help you completing the forms well or which hinder you to do that work? How has the new system changed your cooperation with other teams outside the department, e.g. your supervisors, the HMT, the CHMT? Which members of these groups do you talk more often to, to which of them less? How have the topics of your communication or cooperation have changed? How have you integrated the new SPT forms in your daily work? How have the new forms or the work you do with them changed your daily routine? How has the use of the new forms changed the way you report to vertical programmes, such as HIV?)*

2 A. **Can you share any experiences of times when you have *used* health information for mothers and newborns *after* the introduction of the new system?**

*(Prompts: How do you think data can be useful to improve patient care and your work? Which effect do you think the data you get from the forms have on your work and on patient care? How do you perceive the usefulness of the new system with regards to data quality, availability of data in comparison to HMIS and DHIS? Which changes in the use of data have you noticed at your own department? How do your superiors/facility management/CHMT support you in suing data for quality improvement? Has this changed after the introduction of SPT forms? For what purposes are data used at facility level/CHMT level/national level? How useful your contribution to these uses of data? Please provide examples from your use of data in the last two months: In what situation? For what purpose? For Quality improvement? Has the way you have used health information changed? Have you noticed differences in the way you discussed data and made decisions about quality improvement/planning/care provision priorities? Please give examples)*

2 B. **How do you feel about the *availability* of information (including feed-back from the district) following the introduction of the SPT system?** *(Prompts: Is information more readily available at facility level? How do you feel about the level of correctness and completeness of the information you get? Do you see any changes or is this the same?)*

3 A. **Can you give examples of any *benefits for your work* that you have experienced with the SPT system compared to the regular HMIS system?**

*(Prompts: usefulness of data, timely feed-back, benefits with regards to patient care, time spent on documentation, priority setting, decision making in quality improvement? Which parts of the new process and which parts of the forms do you find most useful in their contribution to facilitating your work or improving the data? Which parts less so and why?)*

3 B. **Can you give examples of any *challenges* you have experienced with the SPT system compared to the regular HMIS system?**

*(Prompts: difficulty to fill forms, not having monthly summary in facility etc.)*

3C. **Do you feel your knowledge and skills concerning completing the form and data interpretation and use for quality improvement are adequate, why?**

3D. **Which mechanisms do you have at the department or health facility to monitor reporting and data quality after introduction of the SPT technology?**

(Prompts: *Informal ones/Formal ones? Who has introduced this monitoring, the management, in charges or staff themselves? How do you as individuals check if the work is done well and if the new technology has improved data or your work or even patient outcomes?)*

*No 4 where applicable:*

4A. **You mentioned other parallel reporting systems during our first meeting (FGD). Which ones are still in place and have anything changed regarding the reporting into these systems following the introduction of the SPT system?**

**4B.We understand that sometimes similar or the same information is recorded. Do you see any discrepancies in terms of figures between the SPT data and the data you collect for other programmes? Pls give examples.**

#### Annex B: Topic guide In-depth interviews post-implementation (English version)

#### INTERVIEW GUIDE FOR REPEAT INTERVIEWS WITH CHMT MEMBERS AND OTHER KEY INFORMANTS AT DISTRICT LEVEL

##### Respondents

CHMT member and other key informants with knowledge and engagement in health information for maternal and newborn care in intervention districts

##### Objective

To establish the acceptability, adoption, feasibility and appropriateness of the adapted Smart paper Technology ™ system for maternal and newborn care and to evaluate its usefulness for supporting ongoing Quality improvement activities in health facilities

##### INTRODUCTION

*Purpose of the interview*

We are in the process of testing a new and simplified health information system in health facilities in this district. We would therefore like to understand your perceptions of the current health information system (HMIS) – the processes involved in documenting, receiving, managing and analysing data from health facilities and how this information is used at the district level and in health facilities for quality improvement and other activities. We would also like to learn about other /parallel/ reporting systems (other than HMIS for example programme specific systems) that may be in place in this district and later to return to learn about your experiences of the new system once it has been implemented.

*Introduce the interviewer and translator*

*Consent procedure*

*Check-list (same for all three repeat interviews)*

##### **SECTION 2:** SECOND INTERVIEW EARLY ASSESSMENT OF SPT IMPLEMENTATION

##### INTRODUCTION

*Last time we met, we discussed the HMIS system for maternal and newborn health information in the district.* ***(Possibility: Provide a summary of what was discussed during the previous interview; this can serve both as a reminder and starting point of this new interview as well as a way of "validating" and feeding back the results from the first interview)****. Since then, the project has introduced the new system which has changed the processes around documenting, reporting and feed-back of health information. We are now interested to learn about your experiences of this new system and how you perceive it compared to the regular HMIS system, what is good and what is challenging and what changes you have experienced and observed.*

1 A. **What did you think when you first heard about the smart paper technology?**

( Prompts: *How difficult was it to understand? What were your feelings about how health workers would be able to apply the new forms? What did you hope to gain from the new system for your work? Does this still hold or did your perception change after using the new system for one month? Why?*)

1B. **What do health care providers think of the new system?**

(Prompts: *Do they see any benefits for their own work? If yes, which benefits did you hear about? Has their attitude towards use of data for their daily work changed? If yes how?)*

1C**. How did you or the facility management support the introduction of the use of SPT forms?**

(Prompts: *How do you normally introduce a new tool or a new task at health facilities? How about the introduction of SPT forms? Was this any different?* If yes, how? *How did you monitor their use and make sure, that all health care providers know what to do and how? How do you make sure that providers continue to use the forms well*? How did you or the facility management make sure that all health care providers have the necessary knowledge, tools and skills to use the new system?)

1D. **How did you and the facility management make sure that all health care providers are on board and understand the advantages and disadvantages of the new system?**

(Prompts: How did you or the facility management deal with resistance or reluctance to use the forms? What are factors that influence their application of the new system?)

1E. **Can you describe any changes you have experienced or observed in the health information system for maternal and newborn care in the district following the introduction of the SPT system**

*(Prompts: How do you perceive the effects, the new system has on your work as CHMT member, as a data person? Where and when documentation is done, monthly reporting from health facilities to the districts, processing of data, have any challenges you experienced with the regular HMIS been solved?)*

1F. **Can you describe any changes you have experienced specifically with the *monthly reporting* of health information for mothers and newborns from health facilities following the introduction of the SPT system**

*(Prompts: How is it different, who is responsible, time taken, when is it done, have any challenges you experienced with the regular HMIS system been solved? How have you integrated the use of the SPT technology in your daily work? How do you see the fit of the new system within the work processes of the facilities or the CHMT?)*

1G. **How have the new system affected your involvement/activities around the processing of health information for mothers and newborns in the district?**

*(Prompts: change in responsibilities/activities, How has the introduction of the SPT technology shaped your cooperation with facility management, in charges or health care providers? Which members of these groups do you contact more often now, which ones less? How has the content of your communication/cooperation changed since you are using the forms?)*

1H. **How has the new system affected the way that you provide feedback to health facilities in the district?**

(Prompts: Frequency, content, format, context-when/in what situations/for what reason etc.)

2 A. **Can you share experiences of *using* health information for mothers and newborns *after* the introduction of the new system?**

*(Prompts: How do you perceive the usefulness of the new system with regards to the use of data, at your level, at facility level, at departmental level? How do you perceive the availability and quality of data now? Are you more willing to use them for your daily work? If yes, why, if not, why not? Please give examples of use of data from the SPT system you have witnessed in the last two months: In what situation? For what purpose? For*

*Quality improvement? Has the way you have used health information changed? How much do you trust data quality now and before? What do you do to assist HP to make better use of the data they have now?)*

2B. **Do you feel your knowledge and skills concerning using the data derived from the SPT technology are sufficient to make good use of these data?**

(Prompts: *How could you use them? What other knowledge and skills would you need to better use them?)*

2C. **Do you feel the HCP's knowledge and skills for completing the forms and using the data well for decision making or quality improvement are adequate?**

(Prompts: *Which additional skills or knowledge would they need to make best use of the data?)*

2 D. **How did the processes required to *access and use* health information for mothers and newborns compare with those of the HMIS system?**

*(Prompts: ease of access, computerized data etc.)*

2 E. **How do you feel about the *availability* of information following the introduction of the SPT system?**

*(Prompts: timeliness, completeness etc.)*

3 A. **Can you give examples of any *benefits* you have experienced with the SPT system compared to the regular HMIS system?**

*(Prompts: How do you rate the usefulness of the SPT system for your own work and for HCPs work in comparison to HMIS and DHIS? How do you rate the quality of reports and tally sheets? How is their timeliness and how did these affect your work, especially concerning your Data Quality Assurance and the reporting to different actors, e.g. regional medical office, MoHCDGEC, vertical programmes? Which advantages do you see for your work and the work of health care providers?)*

3 B. **Can you give examples of any *challenges* you have experienced with the SPT system compared to the regular HMIS system?**

*(Prompts: issues with monthly reporting, scanning station problems, electricity shortages, delays, access to data through DHIS-2)*

3C. **What mechanisms do you have at CHMT/ facility management level to monitor effectiveness and utility of the SPT technology?**

(Prompts: *Informal/formal measures, Who has introduced these mechanisms? How do you as individuals check if the work is done well and if the new technology has improved data or your work or even patient outcomes?)*

*No 4 where applicable:*

4. **You mentioned other parallel reporting systems during the first interview. Which ones are still in place and have anything changed regarding the reporting into these systems following the introduction of the SPT system?**

**Annex C: Task list used for programming**

| | | |
|---|---|---|
| categories | 1 | Personal |
| categories | 2 | Administrative (Hospital) |
| categories | 3 | Patient care labor room (direct care) |
| categories | 4 | Direct patient care ANC |
| categories | 5 | indirect patient care ANC |
| categories | 6 | waiting |
| categories | 7 | Walking |
| categories | 8 | Serving OPD patients |
| categories | 9 | PNC patient care |
| categories | 10 | Miscellaneous |
| personaltsk | 1 | Computer: Email or other (eg browsing) personal |
| personaltsk | 2 | Looking/Waiting: Personal |
| personaltsk | 3 | Miscellaneous: Eating/Drinking/Idle |
| personaltsk | 4 | Miscellaneous: On break |
| personaltsk | 5 | Phone: Personal |
| personaltsk | 6 | Miscellaneous: rest room |
| personaltsk | 7 | Talking: Personal |
| hosp_admintsk | 1 | Filling patient record in register |
| hosp_admintsk | 2 | Looking for patient record in register |
| hosp_admintsk | 3 | Entering patient record in register |
| hosp_admintsk | 4 | Retriving patient record in register |
| hosp_admintsk | 5 | Reffiling new card |
| hosp_admintsk | 6 | Preparing Tally sheet |
| hosp_admintsk | 7 | Preparint monthly summary |
| hosp_admintsk | 8 | Preparing other report (eg PMTCT) |
| hosp_admintsk | 9 | SPT:Filling registration form |
| hosp_admintsk | 10 | SPT:Filling registration update form |
| hosp_admintsk | 11 | SPT:Filling visit form (PNC ANC Labor forms) |
| hosp_admintsk | 12 | SPT:Looking for a record in electronic register |
| hosp_admintsk | 13 | SPT:Reading for a record in electronic register |
| hosp_admintsk | 14 | SPT:Retrieving record in electronic register |

| | | |
|---|---|---|
| hosp_admintsk | 15 | SPT:Filling a new card form electrionic register |
| hosp_admintsk | 16 | SPT:Preparing other monthly summaries (NOT ANC PNC LABOR) |
| hosp_admintsk | 17 | Computer (HC only): |
| hosp_admintsk | 18 | Computer: (HC only): GoTHOMIS data entry |
| hosp_admintsk | 19 | Reading administrative reports |
| hosp_admintsk | 20 | Other work related |
| hosp_admintsk | 21 | Talking to colleague personal |
| hosp_admintsk | 22 | Talking to community health workers on QUADS |
| hosp_admintsk | 23 | Talking to other professionals |
| hosp_admintsk | 24 | Talking to other professionals in person |
| hosp_admintsk | 25 | Filling other forms NOT ANC/Labor/PNC |
| direct_care_labor | 1 | Pelvic examination |
| direct_care_labor | 2 | Taking vital signs |
| direct_care_labor | 3 | Inserting cannula |
| direct_care_labor | 4 | Setting up drip |
| direct_care_labor | 5 | Abdominal examination |
| direct_care_labor | 6 | Talking to patient |
| direct_care_labor | 7 | Talking to family |
| direct_care_labor | 8 | Talking to patient and family or other accompanying person |
| direct_care_labor | 9 | Taking blood |
| direct_care_labor | 10 | Preparing delivery kit |
| direct_care_labor | 11 | Performing vaginal delivery |
| direct_care_labor | 12 | Preparing vacuum delivery |
| direct_care_labor | 13 | Performing vacuum delivery |
| direct_care_labor | 14 | Preparing caesarean section |
| direct_care_labor | 15 | Performing caesarean section |
| direct_care_labor | 16 | Conducting third stage of labour |
| direct_care_labor | 17 | Cleaning patient |
| direct_care_labor | 18 | Assisting with breast feeding |
| direct_care_labor | 19 | Checking fetal heart rate |
| direct_care_labor | 20 | Inserting urinary catheter |
| direct_care_labor | 21 | Checking urine for protein |
| direct_care_labor | 22 | Checking oxygen saturation |
| direct_care_labor | 23 | Caring for newborn |
| direct_care_labor | 24 | Resuscitating newborn |
| direct_care_labor | 25 | Preparing other direct care interventions (giving injections, |

| | | |
|---|---|---|
| | | inserting catheter, inserting iv line) |
| direct_care_labor | 26 | Giving injection im |
| direct_care_labor | 27 | Inserting iv line |
| direct_care_labor | 28 | Giving injection iv |
| direct_care_labor | 29 | Looking for equipment |
| direct_care_labor | 30 | Stitching |
| direct_care_labor | 31 | Dressing and remove stitches |
| direct_care_ANC | 1 | Taking vital signs |
| direct_care_ANC | 2 | Talking to patient |
| direct_care_ANC | 3 | Talking to husband |
| direct_care_ANC | 4 | Talking to patient and husband or other accompanying person |
| direct_care_ANC | 5 | Talking to other accompanying person |
| direct_care_ANC | 6 | HIV/ FP counselling |
| direct_care_ANC | 7 | Taking blood |
| direct_care_ANC | 8 | Doing pelvic examination |
| direct_care_ANC | 9 | Doing abdominal examination |
| direct_care_ANC | 10 | Checking fetal heart rate |
| direct_care_ANC | 11 | Checking urine for protein |
| direct_care_ANC | 12 | Patient education (group) |
| direct_care_ANC | 13 | Measuring weight |
| direct_care_ANC | 14 | Measuring height |
| direct_care_ANC | 15 | Dispensing drugs |
| direct_care_ANC | 16 | Speculum examination |
| direct_care_ANC | 17 | General physical examination |
| direct_care_ANC | 18 | Looking for equipment (e.g. blood pressure cuff) |
| direct_care_ANC | 19 | Providing/inserting fp method |
| direct_care_ANC | 20 | Remving Implant/IUCD |
| indirect_care | 1 | Reading: Patient chart/card |
| indirect_care | 2 | Reading: medical guidelines |
| indirect_care | 3 | Writing: Chart notes |
| indirect_care | 4 | Writing: Orders or prescriptions |
| indirect_care | 5 | Writing: Filling partograph |
| indirect_care | 6 | Writing: Issuing ANC card |
| indirect_care | 7 | Writing: Updating information on ANC card |
| indirect_care | 8 | Talking: Discussing patient with colleagues for decision making |
| indirect_care | 9 | Writing: Referral form/letter |

| | | |
|---|---|---|
| indirect_care | 10 | Phone: Organizing referral |
| indirect_care | 11 | Cleaning labour room post delivery |
| indirect_care | 12 | Order lab examinations |
| indirect_care | 13 | Review lab results |
| indirect_care | 14 | Writing: documenting delivery |
| indirect_care | 15 | Searching: Lab results |
| indirect_care | 16 | Analysing partograph |
| indirect_care | 17 | Decontamination |
| waiting | 1 | Waiting for patient |
| waiting | 2 | Waiting for computer |
| waiting | 3 | Waiting for lab result |
| waiting | 4 | Waiting for colleague |
| waiting | 5 | Waiting for phone call |
| waiting | 6 | Waiting for other work related |
| waiting | 7 | Waiting for Not sure |
| walking | 1 | Walking inside labour room |
| walking | 2 | Walking Outside labour room |
| walking | 3 | Walking Inside RCH |
| walking | 4 | Walking Outside RCH |
| walking | 3 | Walking Inside PNC |
| walking | 4 | Walking Outside PNC |
| walking | 5 | Walking - Bringing specimen to lab |
| walking | 6 | Walking -Looking for equipment |
| miscelaniuos | 1 | Miscellaneous: Cannot find entry |
| miscelaniuos | 2 | Miscellaneous: Observer personal hygiene |
| miscelaniuos | 3 | Miscellaneous: Work related |
| miscelaniuos | 4 | Phone: not sure |
| miscelaniuos | 5 | Read: Non-medical |
| miscelaniuos | 6 | Talking: Not sure |
| miscelaniuos | 7 | Walking inside or outside other than RCH or labour room |
| miscelaniuos | 8 | Staff shift handover |
| gender | 1 | Male |
| gender | 2 | Female |
| cadre | 1 | EN |
| cadre | 2 | CO |

| | | |
|---|---|---|
| cadre | 3 | AMO |
| cadre | 4 | ANO |
| cadre | 5 | MD |
| cadre | 6 | MA |
| cadre | 7 | ACO |
| cadre | 8 | Other |
| pnc_tasks | 1 | Taking vital signs |
| pnc_tasks | 2 | Physical examination of patient |
| pnc_tasks | 3 | Assisting with breast feeding |
| pnc_tasks | 4 | Preparing other direct care interventions (giving injections, inserting/removing catheter, inserting iv line, taking blood) |
| pnc_tasks | 5 | Looking for equipment |
| pnc_tasks | 6 | Talking to patient |
| pnc_tasks | 7 | Talking to family |
| pnc_tasks | 8 | Talking to patient and family or other accompanying person |
| pnc_tasks | 9 | Checking urine for protein |
| pnc_tasks | 10 | Newborn: Physical examination |
| pnc_tasks | 11 | Newborn: Cord care |
| pnc_tasks | 12 | Newborn: Vital signs |
| pnc_tasks | 13 | Newborn: Take blood |
| pnc_tasks | 14 | Newborn: Inserting iv line |
| pnc_tasks | 15 | Newborn: Injecting drugs |
| pnc_tasks | 16 | Newborn: Vaccination (BCG, OPV0) |
| pnc_tasks | 17 | Newborn: Weighing |
| pnc_tasks | 18 | Newborn: Assisting with Kangaroo (HC and dispensary only) |
| pnc_tasks | 19 | Dressing and remove stitches |
| pnc_tasks | 20 | councelling on (BF/care fo the new born/FP) |
| | | removing catheter/ drip |
